# Clear plastic drapes may be effective at limiting aerosolization and droplet spray during extubation: implications for COVID-19

**DOI:** 10.1101/2020.03.26.20044404

**Authors:** C.T. Matava, J. Yu, S. Denning

## Abstract

Protection of frontline HCPs is paramount. However, PPE is a limited resource and often requires providers to be adaptive and resourceful in a crisis. The inexpensive and simple method of using clear drapes during extubation (and possibly intubation) of COVID-19 patients may be considered by frontline HCPs and infection control specialists as an additional precaution. Modifications of the clear plastics can be adapted for surgical procedures that may be AGMPs.

---

To the Editor,

Health care providers (HCPs) performing aerosol-generating medical procedures (AGMP) in patients with the disease caused by severe acute respiratory syndrome coronavirus 2 (SARS-CoV-2), also known as coronavirus disease (COVID)-19, are considered to be at higher risk for contracting the disease.^1^ Reports on the airborne transmission of SARS-CoV-2 have resulted in concerns over increased risk for infection, especially with global shortages of personal protection equipment (PPE) needed for droplet precautions.^2^ Contamination of surfaces and personnel with virus-loaded droplets may occur during high-risk aerosol-generating medical procedures (AGMP), such as intubation and extubation. A recent report recommended the use of gauze around the patient’s mouth as a method for reducing the aerosolization of the virus.^1^ Our group performed a series of experiments assessing whether clear plastic drapes were effective in containing aerosolization during extubation. These experiments did not require institutional research ethics approval.

All experiments utilized Glo-germ™ (Glo Germ Company, Moab, UT, USA), a fluorescent resin powder with particle sizes between 1-5 µm (SARS-COV-2 is 0.07-1.2 µm), with ultraviolet light detection in a darkened operating room. A pediatric manikin (Eletripod ET/J10 Tracheal Intubation model, Tuqi, China) was used wherein simulated secretions (0.5 mL of Glo-germ) were applied to the oropharynx and the mid-trachea of the manikin. A cough was simulated using a calibrated medical air gun connected to the distal trachea, fired over 0.4 sec, delivering cough peak expiratory flow rates (CPEFR) of 150-180 L·min^-1^ outward from the trachea. The normal CPEFR ranges from 87 L·min^-1^ in children under one year, to 728 L·min^-1^ in some adults; a **CPEFR** < 175 L·min^-1^ is predictive of failure to extubate in adults.^**3-5**^ Between experiments, the manikin was cleaned with alcohol, soap and water. All experiments were video recorded (240 fps) using a combination of GoPro Hero 6 (GoPro Inc, San Mateo, CA, USA), iPhone X, X Max, and XR (Apple Inc, Cupertino, CA, USA).

In the first series of experiments (Exp), we simulated a cough during extubation of the manikin both without (Exp 1A) and with (Exp 1B) a single clear plastic drape applied over the head and endotracheal tube. We then repeated the same extubation cough sequence in a second series of experiments (Exp 2) using a modified three-layer plastic drape configuration. The first layer was placed under the head of the manikin to protect the operating table and linen. The second torso-drape layer was applied from the neck down and over the chest, preventing contamination of the upper torso. The final over-head top drape was a clear plastic drape with a sticky edge that was secured at the mid sternum level. It was draped over the patient’s head to prevent contamination of the surrounding surfaces, including the HCPs. After the cough was complete, the top two drapes were rolled away together toward the patient’s legs, to contain and remove the contaminant and the third drape removed afterward.

A cough without any plastic drapes applied (Exp 1A) resulted in a wide distribution of droplets contaminating the surrounding areas (Figure A and eVideo in the Electronic Supplementary Material [ESM]).^A^ The use of a single clear plastic drape (Exp 1B) restricted the aerosolization and droplet spraying of the particles, but using the three-drape technique (Exp 2) resulted in significant further reduction in contamination of the immediate area surrounding the patient (Figure B; eVideo in the ESM). An area of significant contamination, a “hot-zone” was noted on the drape covering the bed beneath the mannequin head (Figure C). The patient’s face and head were also contaminated (Figure C). However, we were able to successfully remove these drapes (by rolling them up) without further contamination of the area or the HCP. This was evidenced by the lack of fluorescent particles on visual inspection.

Our series of experiments (each performed once) were proof-of-concept on the patterns of aerosolization and droplet sprays during extubation and the impact of clear drapes. We were to demonstrate that the use of low-cost barriers (clear plastic drapes) were able to significantly limit the aerosolization and droplet spray. Protection of frontline HCPs is paramount. However, PPE is a limited resource and often requires providers to be adaptive and resourceful in a crisis. The inexpensive and simple method of using clear drapes during extubation (and possibly intubation) of COVID-19 patients may be considered by frontline HCPs and infection control specialists as an additional precaution. Modifications of the clear plastics can be adapted for surgical procedures that may be AGMPs. Limitations of this work include its low-fidelity design and use of a larger particle (Glo-Germ) that may not reflect true spread of a virus like SARS-CoV-2. It is particularly important that HCPs take care not generate further aerosols during removal of the drapes; and recommend using the three-panel draping as opposed to the single drape technique. Further studies will be needed to further refine this model and its findings.

## Data Availability

All data are included in this paper

Clyde T. Matava MBCHB MMed MHSC

Julie Yu MD FRCA

Simon Denning BMBS BMedSci FRCA

Department of Anesthesia and Pain Medicine, The Hospital for Sick Children; Department of Anesthesia, Faculty of Medicine, University of Toronto, Toronto, ON, Canada

; Twitter: @innov8doc

## Funding statement

None.

This submission was handled by Dr. Hilary P. Grocott, Editor-in-Chief, *Canadian Journal of Anesthesia*.

## Footnote

A. This concept has been reported elsewhere. Available from URL: https://twitter.com/innov8doc/status/1240455223929458696 (accessed March 2020).

---

**FIGURE.**
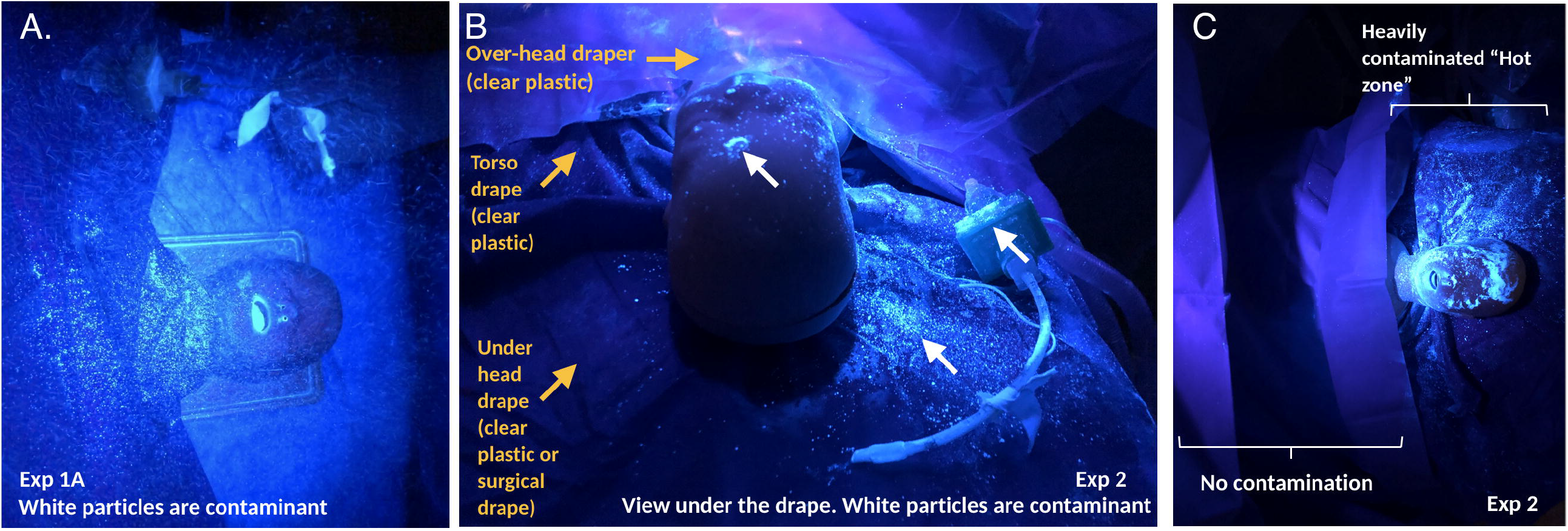
The distribution of particles following the use of a three-layered clear plastic drape configuration for extubation with a simulated cough. A) In experiment (Exp) 1A described in the text, coughing during extubation contaminated the airspace around the patient, including the torso, face, head. and bed B) In Exp 2, a three-panel clear plastic drape was used: first layer placed under the head of the manikin to protect the operating table and linen; second torso-drape layer applied from the neck down and over the chest, preventing contamination of the upper torso; third over-head top drape with a sticky edge secured at the mid sternum level. The clear plastic drapes restrict contamination (white fluorescent particles) to the areas between the top clear plastic and the bottom clear plastic drape, as seen in the view from the patient’s head under the third drape covering the face. C) Following removal of the top clear plastic drape and torso clear plastic drape in Exp 2, the contamination is restricted to the area previously under the top clear plastic drape. There is no contamination of the area previously covered by the torso clear plastic,

## Electronic Supplementary Material

eVIDEO The use of a single clear plastic drape in minimizing aerosolization, droplet spraying, and room contamination. This concept has been reported elsewhere (https://twitter.com/innov8doc/status/1240455223929458696)

